# Discovery of 318 novel loci for type-2 diabetes and related micro- and macrovascular outcomes among 1.4 million participants in a multi-ethnic meta-analysis

**DOI:** 10.1101/19012690

**Authors:** Marijana Vujkovic, Jacob M. Keaton, Julie A. Lynch, Donald R. Miller, Jin Zhou, Catherine Tcheandjieu, Jennifer E. Huffman, Themistocles L. Assimes, Renae L. Judy, Jie Huang, Kyung Min Lee, Derek Klarin, Saiju Pyarajan, John Danesh, Olle Melander, Asif Rasheed, Nadeem Qamar, Saqib S. Sheikh, Shahid Hameed, Irshad H. Qureshi, Muhammad N. Afzal, Uzma Jahazaib, Anjum Jalal, Shahid Abbas, Xin Sheng, Long Gao, Klaus H. Kaestner, Katalin Susztak, Yan V. Sun, Scott L. Duvall, Kelly Cho, Jennifer S. Lee, John M. Gaziano, Lawrence S. Philips, James B. Meigs, Peter D. Reaven, Peter W. Wilson, Todd L. Edwards, Daniel J. Rader, Scott M. Damrauer, Christopher J. O’Donnell, Philip S. Tsao, HPAP Consortium, Regeneron Genetics Center, VA Million Veteran Program, Kyong-Mi Chang, Benjamin F. Voight, Danish Saleheen

## Abstract

We investigated type 2 diabetes (T2D) genetic susceptibility in a multi-ethnic meta-analysis of 228,499 cases and 1,178,783 controls in the Million Veteran Program (MVP) and other biobanks. We identified 558 autosomal and 10 X-chromosome T2D-associated variants, of which 286 autosomal and 7 X-chromosome variants were previously unreported. Ancestry-specific analyses identified 25 additional novel T2D-susceptibility variants. Transcriptome-wide association analysis detected 3,568 T2D-associations with T2D-colocalized genetically predicted gene expression of 804 genes in 52 tissues, of which 687 are novel. Fifty-four of these genes are known to interact with FDA-approved drugs and chemical compounds. T2D polygenic risk score was strongly associated with increased the risk of T2D-related retinopathy, and additionally showed evidence for association with chronic kidney disease (CKD), neuropathy, and peripheral artery disease (PAD). We investigated the genetic etiology of T2D-related vascular outcomes in the MVP and observed statistical SNP-T2D interactions at 13 variants, including 3 for coronary heart disease, 1 for PAD, 2 for stroke, 4 for retinopathy, 2 for CKD, and 1 for neuropathy. Our findings may identify potential novel therapeutic targets for T2D and genomic pathways that link T2D and its vascular outcomes.

---

Type 2 diabetes mellitus (T2D) - a leading cause of morbidity globally - is projected to affect up to 629 million people by 2045^1^. People with T2D are at increased risk of developing a wide range of macro- and microvascular outcomes^2^, and there are large disparities in prevalence, severity and co-morbidities across global populations. T2D is highly polygenic and over 400 common variants have been identified that confer disease susceptibility; however, most studies have been performed in European and Asian ancestry populations, and dominant risk alleles in other ethnic groups remain to be discovered. The genetic etiology of T2D-related sequelae remain poorly understood. Identifying genetic risks and critical genes for T2D-related micro- and macrovascular disease could inform clinical management strategies, including patient stratification and optimizing study design of randomized controlled trials for novel treatments. The lack of large-scale multi-ethnic deeply phenotyped cohorts linked to genetic data has made it difficult to address these questions.

We conducted a multi-ethnic association study of T2D risk comprised of 228,499 T2D cases and 1,178,783 controls of European, African American, Hispanic, South Asian, and East Asian ancestry. We investigated the association of a T2D polygenic risk score with 3 major T2D-related macrovascular outcomes (coronary heart disease [CHD], ischemic stroke, and peripheral artery disease [PAD]) and 3 microvascular disease (chronic kidney disease [CKD], retinopathy and neuropathy) in the Million Veteran Program (MVP)^3^. We conducted a genome-wide SNP-T2D interaction analysis on vascular outcomes in MVP to identify genetic variants where the effect of SNP on the vascular outcome depends on the context of T2D presence. We also performed association analyses of genetically-predicted expression levels and expression quantitative trait-T2D colocalization analyses to identify the effects of gene-tissue pairs that influence T2D risk through inter-individual variation in expression.

This study differs from prior GWAS of T2D risk by leveraging large-scale clinical data in conjunction with polygenic scores, extensive evaluation of context specificity for genetic effects on T2D vascular sequelae and describing the regulatory circuits that influence T2D risk. These observations move the study of T2D genetics beyond lists of variants and provide novel biological insights with translational potential.

## Results

### Study Populations

We performed a genome-wide, multi-ethnic T2D-association analysis (228,499 cases and 1,178,783 controls) encompassing five ancestral groups (Europeans, African Americans, Hispanics, South Asians and East Asians) by meta-analyzing genome-wide association study (GWAS) summary statistics derived from the Million Veteran Program (MVP)^3^ and other studies with non-overlapping participants: DIAMANTE Consortium^4^, Penn Medicine Biobank^5^, Pakistan Genomic Resource^6^, Biobank Japan^7^, Malmö Diet and Cancer Study^8^, Medstar^9^, and PennCath^9^ (Online Methods, Supplementary Table 1 and 3). MVP participants (n = 273,409) are comprised predominantly of male subjects (91.6%) and were classified as Europeans (72.1%), African Americans (19.5%), Hispanics (7.5%), and Asians (0.9%)(Supplemental Table 2).

**Table 1:** T2D locus discovery in African Americans

| Description | leadSNP | rsid | EA | NEA | EAF | Beta | SE | P | N | Ncojo | literatureSNP |
| --- | --- | --- | --- | --- | --- | --- | --- | --- | --- | --- | --- |
| Novel AA | chr12:38710523 | rs7315028 | G | A | 0.882 | 0.124 | 0.022 | 1.5E-08 | 56150 | 1 | - |
|  | chr12:57968738 | rs11172254 | G | A | 0.817 | 0.097 | 0.017 | 1.8E-08 | 56150 | 1 | - |
|  | chr12:88338461 | rs10745460 | T | A | 0.660 | 0.079 | 0.014 | 3.7E-08 | 56150 | 0 | - |
| Novel TE | chr7:50887174 | rs7781440 | C | T | 0.284 | -0.086 | 0.015 | 5.3E-09 | 56150 | 0 | - |
|  | chr12:80985872 | rs1528287 | G | T | 0.059 | -0.494 | 0.080 | 8.2E-10 | 56150 | 1 | - |
| Established | chr3:123065778 | rs11708067 | G | A | 0.151 | -0.118 | 0.018 | 2.3E-11 | 56150 | 0 | chr3:123082398 |
|  | chr3:185534482 | rs9859406 | G | A | 0.257 | -0.115 | 0.015 | 5.7E-14 | 56150 | 0 | chr3:185829891 |
|  | chr5:55807370 | rs464605 | C | T | 0.429 | -0.077 | 0.013 | 1.1E-09 | 56150 | 0 | chr5:55860781 |
|  | chr6:39016636 | rs10305420 | C | T | 0.920 | 0.142 | 0.025 | 8.5E-09 | 56150 | 0 | chr6:39282371 |
|  | chr7:15064896 | - | G | T | 0.565 | 0.101 | 0.013 | 2.7E-15 | 56150 | 0 | chr7:15060429 |
|  | chr7:28180556 | rs864745 | C | T | 0.257 | -0.083 | 0.014 | 1.1E-08 | 56150 | 0 | chr7:28198677 |
|  | chr7:44185088 | rs2908274 | G | A | 0.359 | -0.089 | 0.014 | 5.4E-11 | 56150 | 1 | chr7:44266184 |
|  | chr8:41510260 | rs12550613 | G | C | 0.310 | -0.114 | 0.014 | 5.5E-16 | 56150 | 0 | chr8:41537318 |
|  | chr8:118166327 | rs60461843 | T | A | 0.939 | 0.172 | 0.028 | 1.3E-09 | 56150 | 1 | chr8:118024315 |
|  | chr9:139241595 | rs28562046 | G | C | 0.709 | 0.080 | 0.014 | 2.8E-08 | 56150 | 0 | chr9:139737088 |
|  | chr10:114758349 | rs7903146 | C | T | 0.706 | -0.226 | 0.014 | 5.6E-60 | 56150 | 0 | chr10:114871594 |
|  | chr11:2691500 | rs231361 | G | A | 0.656 | -0.080 | 0.013 | 2.2E-09 | 56150 | 2 | chr11:2717680 |
|  | chr11:2858546 | rs2237897 | C | T | 0.908 | 0.143 | 0.024 | 2.2E-09 | 56150 | 1 | chr11:2717680 |
|  | chr12:66215214 | rs2583938 | T | A | 0.197 | -0.123 | 0.018 | 3.3E-12 | 56150 | 0 | chr12:66358347 |
|  | chr15:77776498 | rs952471 | G | C | 0.534 | 0.077 | 0.013 | 4.2E-09 | 56150 | 0 | chr15:77339496 |
|  | chr16:53811788 | rs62033400 | G | A | 0.102 | 0.151 | 0.021 | 6.1E-13 | 56150 | 1 | chr16:53758720 |

### Single-variant autosomal analyses

We identified a total of 558 independent sentinel SNPs (286 novel, 272 previously reported)^4,7,10,11^ associated with T2D (Figure 1, Table 1, Supplemental Tables 4, 5, 6, 10, and 11, Supplemental Figure 1a-e). Variants were classified as novel if they were at least 500 kb distant from a previously reported sentinel SNP^4,7,10,11^ and/or if they were not in linkage disequilibrium (LD, r^2^ ≤ 0.05) with an established sentinel SNP^4,7,10,11^. A total of 21 SNPs showed genome-wide significance in Europeans only (Supplemental Table 6). We found that novel loci had, on average, smaller magnitudes of effect (0.032 ± 0.012 per allele) than previously established SNPs (0.054 ± 0.045 per allele, derived from data shown in Supplementary Table 5), presumably resulting from enhanced power to discover weaker effects due to the large sample size and ancestral diversity. Genome-wide chip heritability analysis explained 19% of T2D risk^4^.

**Figure 1:**
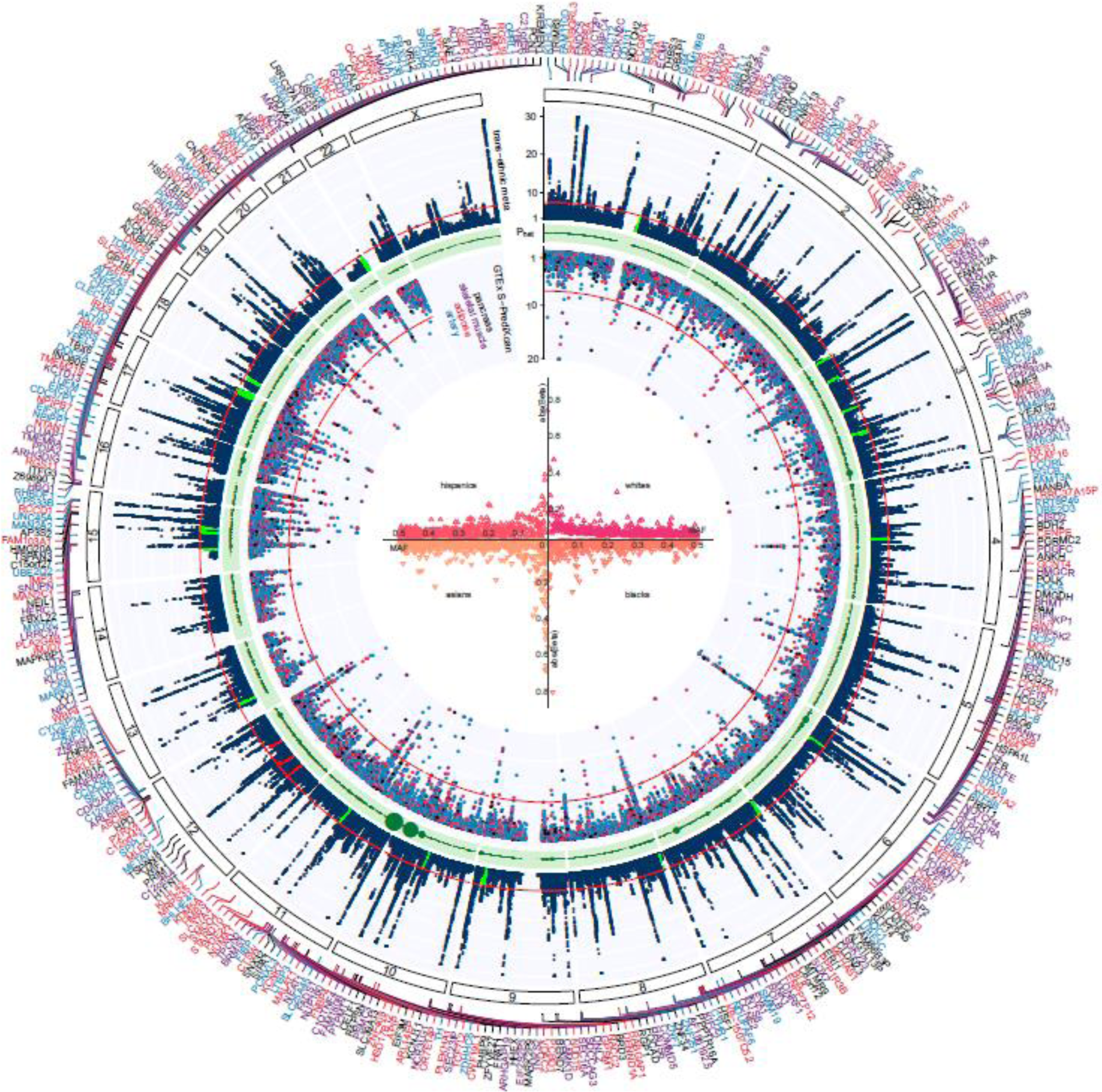
Trans-ancestry GWAS meta-analysis identifies 318 loci associated with T2D. The graph represents a circos plot. The outer track corresponds to –log10 (P) for association with T2D in the trans-ethnic meta-analysis (y axis truncated at 30), by chromosomal position. The red line indicates genome-wide significance (P=5.0×10−08). Purple gene labels indicated genes identified in skeletal muscle eQTLs by S-PrediXcan analysis, red-labeled gene names in adipose eQTLs, black-labeled gene names in pancreas eQTLs, and blue-labeled gene names were identified in eQTLs from arteries. The green band corresponds to measures of heterogeneity related to the index SNPs associated with T2D. Dot sizes are proportional to I2 or ancestry-related heterogeneity. The inner track corresponds to – log10(P) for association with skeletal muscle, adipose, pancreas, and artery tissue eQTLs from S-PrediXcan analysis (y axis truncated at 20), by chromosomal position. The red line indicates genome-wide significance (P=5.0×10−08). Inset, effects of all 318 index SNPs on T2D by minor allele frequency, stratified and colored by ancestral group.

In an analysis of African American participants (Table 1), we observed a total of 21 loci associated with T2D susceptibility at genome-wide significance, 16 of which were in strong LD with established T2D variants. Three variants were novel and the effect on T2D appeared specific to African-Americans. The frequency of the T2D risk-increasing allele is notably higher in African reference populations than European for these SNPs, with the ancestral allele the major allele in African populations and the derived allele being the major allele in European populations. Single variant analysis in the Hispanics subset identified 2 SNPs, both of which tagged previously reported T2D loci (Supplemental Table 10). No novel variants were observed in the subjects of Asian ancestry (Supplemental Table 11).

### Chromosome-X analyses

A total of 10 association signals for T2D were identified on chromosome X in a multi-ethnic meta-analysis, of which 7 were novel (Table 2a-b, Supplemental Figure 2a-e). A European-restricted analysis identified 4 chromosome-X loci associated with T2D risk, all of which were identified in the multi-ethnic meta-analysis. One novel chromosome-X locus was associated with T2D that is specific to African Americans. Of interest is one novel, multi-ethnic locus near the androgen receptor gene that was in strong LD with a previously reported variant (rs4509480) known to be associated with male-pattern baldness (EUR r^2^=0.98, rs200644307).

**Table 2a:**
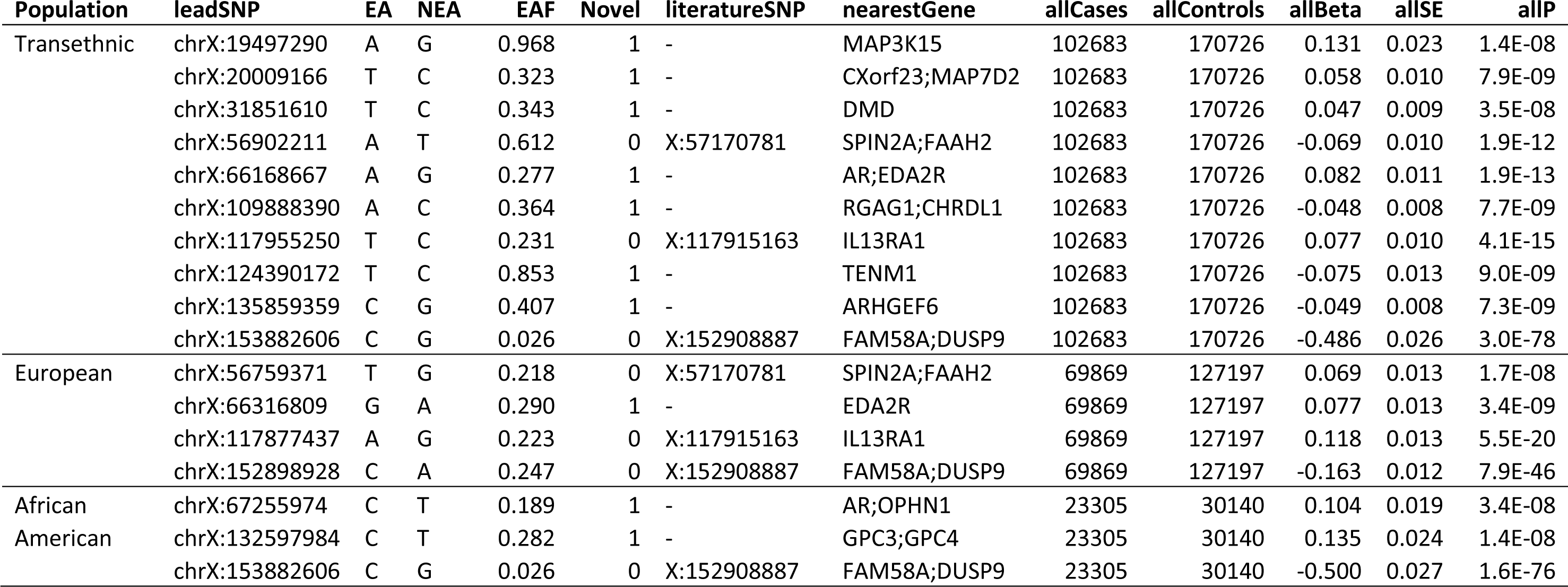
T2D chromosome X analysis (overall results)

**Table 2b:** T2D chromosome X analysis (stratified results by gender)

| Population | leadSNP | EA | EAF | mCase | mCntrl | mBeta | mSE | mP | fCase | fCntrl | fBeta | fSE | fP | hetP |
| --- | --- | --- | --- | --- | --- | --- | --- | --- | --- | --- | --- | --- | --- | --- |
| Transethnic | chrX:19497290 | A | 0.968 | 97456 | 153021 | 0.128 | 0.025 | 5.1E-07 | 5227 | 17705 | 0.147 | 0.056 | 8.0E-03 | 0.751 |
|  | chrX:20009166 | T | 0.323 | 97456 | 153021 | 0.065 | 0.011 | 5.8E-10 | 5227 | 17705 | -0.005 | 0.030 | 8.7E-01 | 0.029 |
|  | chrX:31851610 | T | 0.343 | 97456 | 153021 | 0.051 | 0.009 | 1.3E-08 | 5227 | 17705 | 0.011 | 0.025 | 6.7E-01 | 0.130 |
|  | chrX:56902211 | A | 0.612 | 97456 | 153021 | -0.074 | 0.010 | 1.0E-12 | 5227 | 17705 | -0.028 | 0.031 | 3.6E-01 | 0.156 |
|  | chrX:66168667 | A | 0.277 | 97456 | 153021 | 0.080 | 0.012 | 1.4E-11 | 5227 | 17705 | 0.094 | 0.033 | 4.3E-03 | 0.696 |
|  | chrX:109888390 | A | 0.364 | 97456 | 153021 | -0.044 | 0.009 | 4.2E-07 | 5227 | 17705 | -0.075 | 0.025 | 2.3E-03 | 0.240 |
|  | chrX:117955250 | T | 0.231 | 97456 | 153021 | 0.082 | 0.010 | 2.4E-15 | 5227 | 17705 | 0.034 | 0.029 | 2.5E-01 | 0.118 |
|  | chrX:124390172 | T | 0.853 | 97456 | 153021 | -0.075 | 0.014 | 1.1E-07 | 5227 | 17705 | -0.075 | 0.034 | 2.7E-02 | 1.000 |
|  | chrX:135859359 | C | 0.407 | 97456 | 153021 | -0.052 | 0.009 | 4.8E-09 | 5227 | 17705 | -0.021 | 0.026 | 4.3E-01 | 0.250 |
|  | chrX:153882606 | C | 0.026 | 97456 | 153021 | -0.517 | 0.029 | 1.7E-72 | 5227 | 17705 | -0.349 | 0.061 | 1.1E-08 | 0.013 |
| European | chrX:56759371 | T | 0.218 | 67020 | 115901 | 0.076 | 0.012 | 1.1E-09 | 2849 | 11296 | -0.014 | 0.039 | 7.3E-01 | 0.035 |
|  | chrX:66316809 | G | 0.290 | 67020 | 115901 | 0.073 | 0.014 | 1.1E-07 | 2849 | 11296 | 0.134 | 0.042 | 1.4E-03 | 0.126 |
|  | chrX:117877437 | A | 0.223 | 67020 | 115901 | 0.118 | 0.000 | 3.6E-19 | 2849 | 11296 | 0.121 | 0.041 | 3.1E-03 | 0.943 |
|  | chrX:152898928 | C | 0.247 | 67020 | 115901 | -0.166 | 0.000 | 1.6E-44 | 2849 | 11296 | -0.131 | 0.037 | 4.6E-04 | 0.377 |
| African | chrX:67255974 | C | 0.189 | 21316 | 25287 | 0.117 | 0.020 | 2.6E-09 | 1989 | 4853 | 0.021 | 0.041 | 6.1E-01 | 0.030 |
| American | chrX:132597984 | C | 0.282 | 21316 | 25287 | 0.121 | 0.027 | 7.1E-06 | 1989 | 4853 | 0.231 | 0.059 | 8.9E-05 | 0.080 |
|  | chrX:153882606 | C | 0.026 | 21316 | 25287 | -0.519 | 0.029 | 2.2E-70 | 1989 | 4853 | -0.366 | 0.062 | 4.1E-09 | 0.025 |

**Figure 2:**
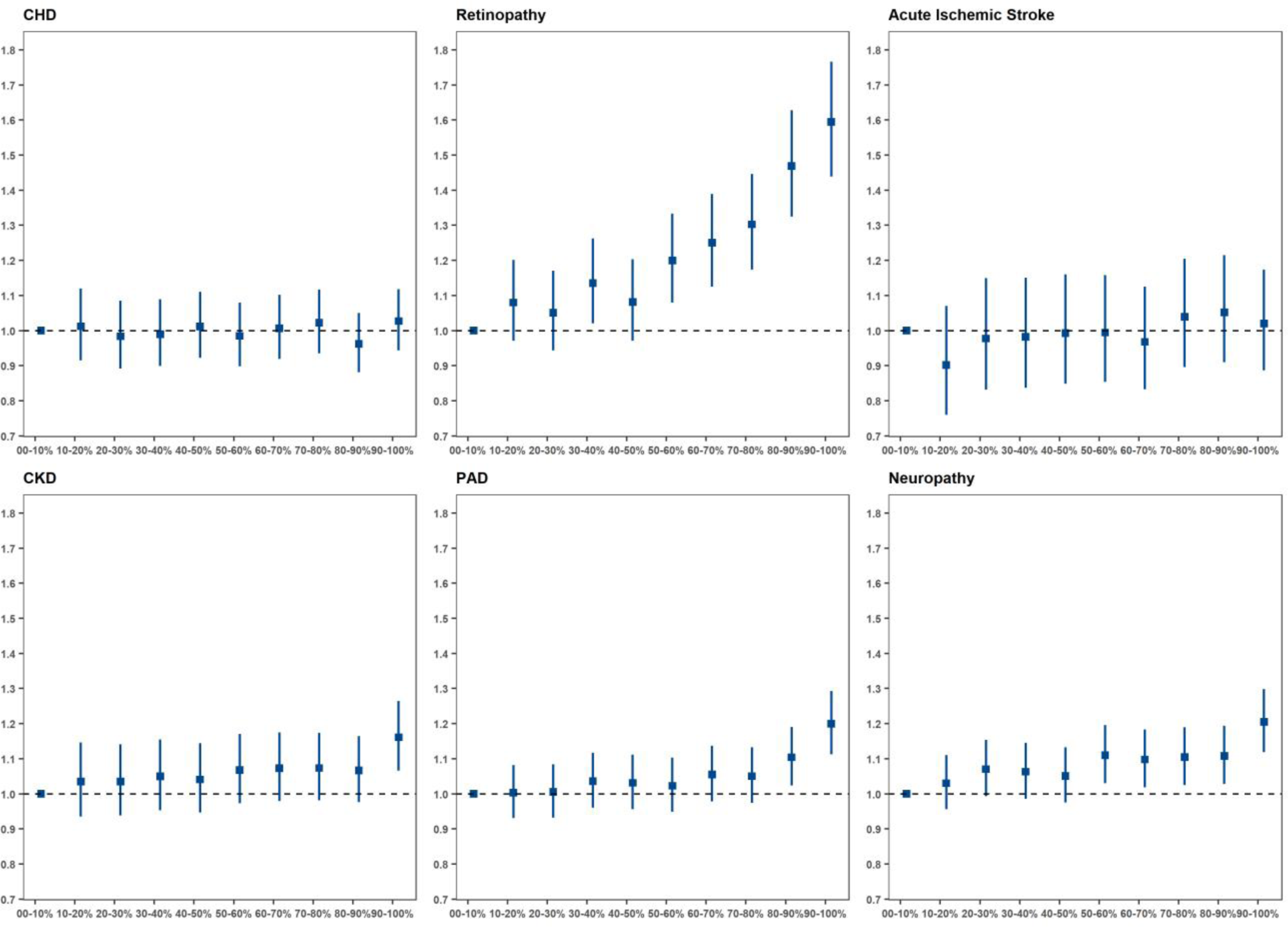
T2D gPRS is mainly predictive of micro vascular outcomes. A T2D gPRS was calculated and categorized into deciles based on the scores in controls. The PRS-outcome associations are shown for macrovascular outcomes (CHD, CKD and PAD) and for microvascular outcomes (retinopathy, neuropathy, and acute ischemic stroke). Effect sizes and 95% confidence intervals are shown per decile per micro- or macrovascular outcome. For each of the complication outcomes separate logistic regression model are fitted for people with T2D, and the models include the following independent variables: T2D PRS (from Diamante consortium), age, gender, BMI, and 10 PCA’s. For coronary heart disease, a CHD PRS (from CardiogramplusC4DplusUKBB) is included in the regression model as an additional covariate. For acute ischemic stroke, a stroke PRS (from Megastroke consortium) is included in the regression model as an additional covariate. For chronic kidney disease, a CKD PRS (from CKDgen consortium) is included in the regression model as an additional covariate.

### Effect heterogeneity between Europeans and African Americans

For most loci, we found no evidence of significant heterogeneity of effect estimates between Europeans and African Americans, whereas 44 (7.9%) had significantly different effect estimates (Supplemental Table 20). Overall, the strength of effect was found to be highest in Europeans, an expected result given the abundance of individuals of that ancestry in our study. However, 4 loci nearby the genes *SLC30A8, PTPRQ, GRB10*, and *COLB* showed higher effect sizes for T2D at stronger levels of significance in African Americans compared with Europeans. Of these loci, associations with loss of function variants in the *SLC30A8* gene were previously reported in Europeans, African Americans and South Asians^12,13^.

### Secondary signal analysis

We detected a total of 233 conditionally independent SNPs that flank 49 novel and 108 previously reported lead SNPs in Europeans (Supplementary Table 7 and 9), and identified 9 independent variants in Europeans and 3 in African Americans flanking 3 novel and 6 previously reported sentinel SNPs. We observed no novel conditionally independent variants in participants of South Asian, East Asian and Hispanic ancestry.

### Rare coding variant mapping

To identify coding variants that may influence T2D risk by changing protein structures, we investigated predicted loss of function (pLoF) and missense variants in LD with the identified T2D lead variants (Supplementary Table 12). We identified 2 pLoF (*LPL* and *ANKDD1B*) and 45 missense variants in 47 genes that were in LD with at-least one of the T2D lead SNPs (r^2^ > 0.5, MVP reference panel in Europeans) and were associated at a P-value < 1.0×10^−4^. We also identified 9 missense variants that were either within 5 Mb of sentinel SNPs or in moderate to low LD (r^2^ < 0.5) and were associated with T2D at a P-value < 1.0×10^−4^. Of the 56 pLoF and missense variants, 14 missense variants were found to be the sentinel T2D SNPs and 19 variants were in LD with novel lead SNPs, and 37 variants were previously reported.

We then performed a genome-wide screen of all pLoF’s (not bound by proximity to sentinel T2D lead variants) and missense variants available for analysis (Supplemental Table 13) and identified one additional pLoF variant in the *CCHCR1* gene, whereas 37 novel missense variants were associated with T2D at a P-value threshold of 5×10^−8^.

#### Rare coding variant PheWAS

To understand biological pleiotropy and underlying mechanisms, we performed a PheWAS of the 3 pLoF variants associated with T2D in MVP participants of European ancestry (Table 5). These loci included *ANKDD1B* p.Trp480*, *CCHCR1* p.Trp78*, and *LPL* p.*474Ser in the MVP, and were significantly associated with metabolic and inflammatory conditions. For example, *ANKDD1B* p.Trp480* was associated with dyslipidemia, hypercholesterolemia, and diabetic neurological manifestations. *CCHCR1* p.Trp78* was associated with type 1 diabetes, epistaxis, celiac disease, microscopic hematuria, and psoriatic arthropathy. Finally, the recent report by Klarin *et al*. includes a Phewas of *LPL* p.*474Ser in MVP^14^, which was associated with dyslipidemia, coronary atherosclerosis and other chronic ischemic heart disease.

#### Transcriptome-wide association analyses

Common variants from the European T2D GWAS meta-analysis were used to evaluate the association of genetically predicted gene expression levels with T2D risk across 52 tissues including kidney and islet cells using S-PrediXcan^15^ (Supplemental Table 8, Supplemental Figure 3). We identified 4,468 statistically significant gene-tissue combination pairs genetically predictive of T2D risk, of which 4,211 transcript eQTLs were in LD (r^2^ > 0.5) with T2D signals. We identified 873 genes in this analysis that would not have been identified by nearest-gene annotation alone. The strongest gene-tissue combination signals were for *NRAP* in the cerebellum and *TCF7L2* in the aortic artery.

We additionally used COLOC to identify the subset of significant genes where there was a high posterior probability that the set of model SNPs in the S-PrediXcan analysis for each gene were both causal for the expression and T2D. This analysis refined the results of the TWAS and excluded some results that might be the consequence of LD between causal SNPs for gene expression and T2D. We detected 3,568 gene-tissue pairs where there was statistically significant association with T2D risk and high posterior probability (P4>0.5) of colocalization. The colocalized and significant results included 804 genes. When comparing the 804 genes to the GWAS catalog mapped and reported genes for all prior studies of diabetes or diabetes complications, 687 had not been identified previously.

Hypergeometric enrichment analysis showed that most enriched gene expression signals were in the following tissues: cervical spinal cord, basal ganglia, glomerular kidney, tibial nerve, transformed fibroblasts, and skeletal muscle (Supplemental Table 15). A cross-section of the putative genes identified through transcriptome-wide analysis and the genes identified from the genome-wide coding variant lookup (Supplemental Table 12) identified the genes: *PCNT* p.Ile539Thr, *NMI* p.Leu16Ser, *TRIM66* p.His322Arg, *STARD3* p.Gln117Arg, and *ZNF641* p.Gln363Pro.

#### Assessment of gene–drug relationships

Of the 991 genes identified in S-PrediXcan analyses, 54 genes have documented interactions with a total of 283 FDA-approved drugs and chemical compounds that do not have an indication for T2D treatment or reported ADE in diabetic patients using the SIDER database of drugs and side effects^16^. Using the Drug-Gene Interaction Database^17^ (DGIdb version 3.0) a total of 322 gene-drug combinations were identified for which it is predicted that they would lower blood glucose based on direction of effect on T2D risk with increasing gene expression and drug action (activator or inhibitor)(Supplemental Table 14). Gene-drug combinations included several established T2D loci including *KCNJ11* targeted by 15 compounds (e.g. sulfonylureas, glinides, and p-glycoprotein inhibitors), *SCNA3* targeted by 57 compounds (e.g. anti-arrythmetics, anti-epileptics), *PIK3CB* targeted by 46 compounds (e.g. cancer drugs), *ACE* targeted by 36 compounds (e.g. angiotensin-converting enzyme [ACE] inhibitors), *HMGCR* targeted by 18 compounds (e.g. HMG-CoA reductase inhibitors), *PIK3C2A* targeted by 15 compounds (anti-cancer drugs), *F2* targeted by 11 compounds (anti-coagulants), and *BLK* targeted by 9 compounds (protein kinase inhibitors).

The gene most significant in S-PrediXcan targeted by a non-antiglycemic medication was *PPARG* in EBV-transformed lymphocytes (Supplemental Table 8). The gene most significant in S-PrediXcan targeted by a drug with report of ADE in diabetic patients was *TH* in non-sun-exposed suprapubic skin. The gene most significant in S-PrediXcan targeted by an established anti-glycemic medication was *KCNJ11* in sun-exposed skin from the lower leg.

#### Pathway and functional enrichment analysis

To explore whether our results recapitulate the pathophysiology of T2D, we performed gene-set enrichment analysis with all the variants using DEPICT (P < 1×10^−5^, Supplemental Table 16). MeSH-based analysis showed that several different adipose tissues and sites were enriched (abdominal subcutaneous fat, abdominal fat, adipose tissue, subcutaneous fat, and white adipose tissue). Finally, DEPICT analysis showed that the most significant gene-set involved the AKT2 subnetwork, lung cancer, the GAB1 signalosome, protein kinase binding, signal transduction, and EGFR signaling (Supplemental Table 17).

#### Genetic correlation between T2D and other phenotypes

Genome-wide genetic correlations of T2D were calculated with a total of 774 complex traits and diseases by comparing allelic effects using LD score regression (Online Methods). A total of 270 significant associations were observed (P < 5.0×10^−8^, Supplementary Table 19). The strongest positive correlations were observed with waist circumference, overall health, BMI, and fat mass of arms, legs, body and trunk, hypertension, coronary artery disease, dyslipidemia, alcohol intake, years of education, wheezing, and cigarette smoking. There was also a strong negative correlation with years of education.

#### T2D-related micro- and macrovascular outcomes

Using a genome-wide approach, we investigated SNP-T2D interaction effects associated with T2D-related vascular outcomes among European-descent MVP participants (P < 5×10^−8^, Online Methods, Table 3a-b). The analysis included a total case count of 67,403 for CKD, 56,285 for CHD, 35,882 for PAD, 11,796 for acute ischemic stroke, 13,881 for retinopathy, and 40,475 for neuropathy. Several genome-wide significant interactions were identified where the genetic associations with T2D-related vascular outcomes were modified by T2D (Table 3a-b). We identified 2 loci for CHD (rs1831733 in 9p21 and rs602633 near *SORT1*) and 1 for CKD (rs34857077 in *UMOD*) for which the difference in the effect estimates between T2D strata was genome-wide significant (P < 5.0×10^−08^) and at least one T2D-stratum was genome-wide significant as well. We additionally identified 1 locus for CHD (rs71039916 near *PDE3A*), 1 for CKD (rs2177223 near *TENM3*), 1 for PAD (rs3104154 in *PTDSS1*), 1 for neuropathy (rs78977169 near *NRP2*), 4 for retinopathy (rs76754787 nearby *GJA8*, rs10733997 in *SVILP2*, rs2255624 near *SLC18A2*, and rs4132670 in *TCF7L2*) and 2 for acute ischemic stroke (rs491203 near *TMEM51*, and rs2134937 near *TRIQK*) that showed a genome-wide significance for difference in effect estimates between the T2D strata and nominal significance (P < 0.001) for at least one T2D stratum.

**Table 3a:**
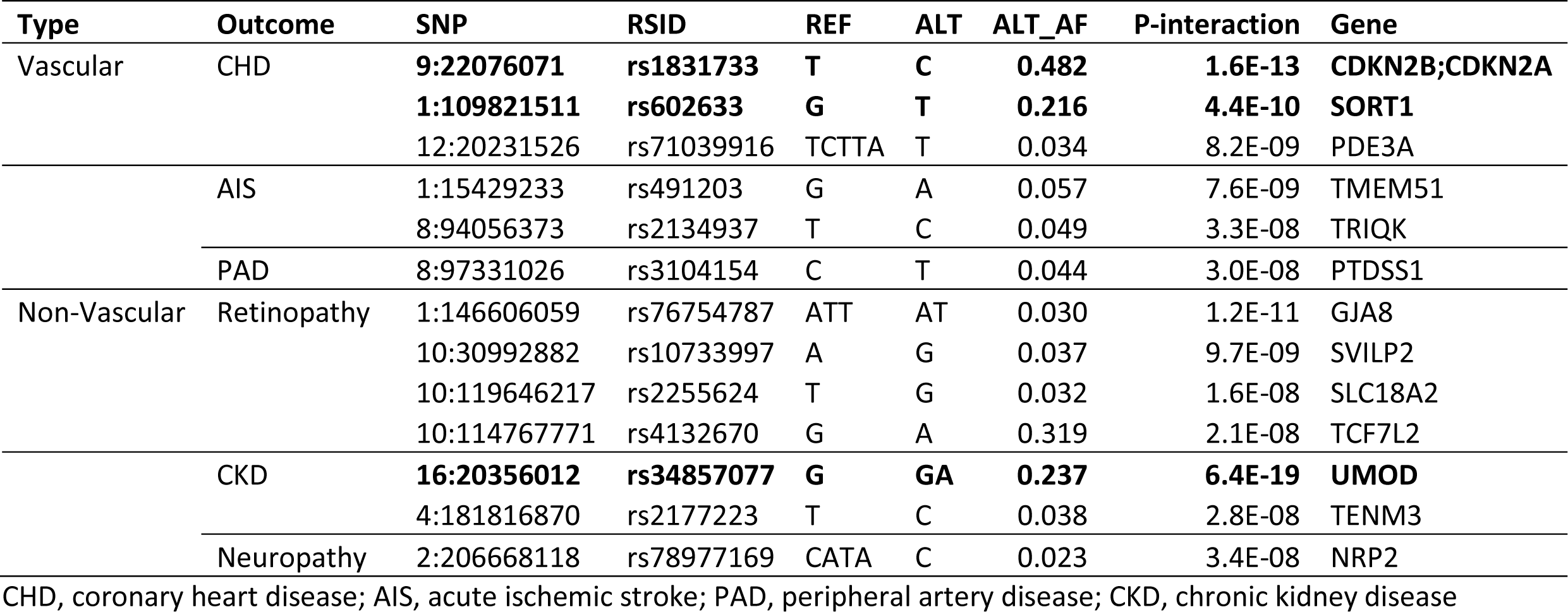
Genome-wide interaction analysis of vascular and non-vascular complications (overall results)

**Table 3b:** Genome-wide interaction analysis of vascular and non-vascular complications (T2D strata specific effect estimates)

| Type | Outcome | rsid | T2D |  |  |  |  | No T2D |  |  |  |  |
| --- | --- | --- | --- | --- | --- | --- | --- | --- | --- | --- | --- | --- |
|  |  |  | Cases | Controls | Beta | SE | P | Cases | Controls | Beta | SE | P |
| Vascular | CHD | **rs1831733** | **30311** | **39568** | **0.123** | **0.011** | **2.6E-29** | **25974** | **101377** | **0.152** | **0.010** | **3.9E-52** |
|  |  | **rs602633** | **30311** | **39568** | **-0.124** | **0.014** | **4.2E-19** | **25974** | **101377** | **-0.108** | **0.013** | **9.9E-17** |
|  |  | rs71039916 | 30311 | 39568 | -0.116 | 0.031 | 9.1E-05 | 25974 | 101377 | -0.002 | 0.029 | 0.945 |
|  | AIS | rs491203 | 5981 | 61240 | -0.161 | 0.044 | 1.3E-04 | 5815 | 117241 | 0.060 | 0.040 | 0.134 |
|  |  | rs2134937 | 5981 | 61240 | -0.153 | 0.047 | 5.7E-04 | 5815 | 117241 | 0.107 | 0.043 | 0.013 |
|  | PAD | rs3104154 | 20223 | 49656 | -0.104 | 0.029 | 1.7E-04 | 15659 | 111692 | 0.044 | 0.029 | 0.129 |
| Non-Vascular | Retinopathy | rs76754787 | 9097 | 44821 | -0.183 | 0.050 | 1.3E-04 | 4784 | 78717 | 0.165 | 0.059 | 0.005 |
|  |  | rs10733997 | 9097 | 44821 | -0.156 | 0.045 | 2.6E-04 | 4784 | 78717 | 0.069 | 0.055 | 0.210 |
|  |  | rs2255624 | 9097 | 44821 | -0.160 | 0.048 | 4.3E-04 | 4784 | 78717 | 0.056 | 0.060 | 0.351 |
|  |  | rs4132670 | 9097 | 44821 | 0.081 | 0.017 | 9.5E-07 | 4784 | 78717 | -0.061 | 0.023 | 0.008 |
|  | CKD | **rs34857077** | **34588** | **35291** | **-0.149** | **0.013** | **1.1E-30** | **32815** | **94536** | **-0.123** | **0.012** | **1.2E-24** |
|  |  | rs2177223 | 34588 | 35291 | -0.117 | 0.030 | 4.8E-05 | 32815 | 94536 | 0.048 | 0.026 | 0.065 |
|  | Neuropathy | rs78977169 | 22943 | 37569 | -0.152 | 0.041 | 1.0E-04 | 17532 | 72762 | 0.041 | 0.040 | 0.305 |

CHD, coronary heart disease; AIS, acute ischemic stroke; PAD, peripheral artery disease; CKD, chronic kidney disease

#### Polygenic risk scores and T2D-related micro- and macrovascular outcomes

Genome-wide polygenic risk scores (gPRS) for T2D were calculated in Europeans based on the T2D effect estimates from the previously reported DIAMANTE consortium^4^ and then categorized into deciles (Table 4a-b). As expected, participants with the highest T2D gPRS scores (90-100% T2D gPRS percentile) showed the highest risk for T2D (OR = 5.21, 95% CI 4.94-5.49, Supplemental Figure 5) when compared to the reference group (0-10% T2D gPRS percentile) in a cross-sectional study design.

**Table 4a:** Polygenic risk scores and vascular complications

| Type | Outcome | T2D PRS-decile | Cases | Controls | OR (95%CI) | P | P-linear* |
| --- | --- | --- | --- | --- | --- | --- | --- |
| Vascular | Coronary Heart Disease<sup>†</sup> | 0-10% | 2913 | 3924 | 1.00 (Ref) | - | 0.636 |
|  |  | 10-20% | 2940 | 3924 | 1.01 (0.92-1.12) | 0.811 |  |
|  |  | 20-30% | 2958 | 3924 | 0.98 (0.89-1.08) | 0.742 |  |
|  |  | 30-40% | 2934 | 3924 | 0.99 (0.90-1.09) | 0.835 |  |
|  |  | 40-50% | 2988 | 3924 | 1.01 (0.92-1.11) | 0.801 |  |
|  |  | 50-60% | 3001 | 3924 | 0.98 (0.90-1.08) | 0.744 |  |
|  |  | 60-70% | 2977 | 3924 | 1.01 (0.92-1.10) | 0.887 |  |
|  |  | 70-80% | 2916 | 3924 | 1.02 (0.93-1.12) | 0.632 |  |
|  |  | 80-90% | 3032 | 3924 | 0.96 (0.88-1.05) | 0.391 |  |
|  |  | 90-100% | 3038 | 3924 | 1.03 (0.94-1.12) | 0.537 |  |
|  | Acute Ischemic Stroke<sup>‡</sup> | 0-10% | 555 | 6027 | 1.00 (Ref) | - | 0.070 |
|  |  | 10-20% | 563 | 6027 | 0.90 (0.76-1.07) | 0.238 |  |
|  |  | 20-30% | 583 | 6027 | 0.98 (0.83-1.15) | 0.782 |  |
|  |  | 30-40% | 619 | 6027 | 0.98 (0.84-1.15) | 0.821 |  |
|  |  | 40-50% | 530 | 6027 | 0.99 (0.85-1.16) | 0.924 |  |
|  |  | 50-60% | 576 | 6027 | 0.99 (0.85-1.16) | 0.941 |  |
|  |  | 60-70% | 645 | 6027 | 0.97 (0.83-1.13) | 0.672 |  |
|  |  | 70-80% | 590 | 6027 | 1.04 (0.90-1.20) | 0.611 |  |
|  |  | 80-90% | 558 | 6027 | 1.05 (0.91-1.22) | 0.494 |  |
|  |  | 90-100% | 627 | 6027 | 1.02 (0.89-1.17) | 0.784 |  |
|  | Peripheral Artery Disease | 0-10% | 1966 | 4871 | 1.00 (Ref) | - | 2.0E-07 |
|  |  | 10-20% | 1964 | 4871 | 1.00 (0.93-1.08) | 0.927 |  |
|  |  | 20-30% | 1948 | 4871 | 1.01 (0.93-1.08) | 0.890 |  |
|  |  | 30-40% | 1984 | 4871 | 1.04 (0.96-1.12) | 0.361 |  |
|  |  | 40-50% | 1964 | 4871 | 1.03 (0.96-1.11) | 0.425 |  |
|  |  | 50-60% | 1950 | 4871 | 1.02 (0.95-1.10) | 0.559 |  |
|  |  | 60-70% | 1972 | 4871 | 1.05 (0.98-1.14) | 0.165 |  |
|  |  | 70-80% | 1960 | 4871 | 1.05 (0.97-1.13) | 0.203 |  |
|  |  | 80-90% | 2019 | 4871 | 1.10 (1.02-1.19) | 0.010 |  |
|  |  | 90-100% | 2102 | 4871 | 1.2 (1.11-1.29) | 1.9E-06 |  |

**Table 4b:** Polygenic risk scores and non-vascular complications

| Type | Outcome | T2D PRS-decile | Cases | Controls | OR (95%CI) | P | P-linear* |
| --- | --- | --- | --- | --- | --- | --- | --- |
| Non-Vascular | Retinopathy | 0-10% | 792 | 4533 | 1.00 (Ref) | - | 3.1E-32 |
|  |  | 10-20% | 832 | 4533 | 1.08 (0.97-1.20) | 0.158 |  |
|  |  | 20-30% | 795 | 4533 | 1.05 (0.94-1.17) | 0.364 |  |
|  |  | 30-40% | 852 | 4533 | 1.14 (1.02-1.26) | 0.019 |  |
|  |  | 40-50% | 814 | 4533 | 1.08 (0.97-1.20) | 0.152 |  |
|  |  | 50-60% | 891 | 4533 | 1.20 (1.08-1.33) | 6.8E-04 |  |
|  |  | 60-70% | 901 | 4533 | 1.25 (1.13-1.39) | 3.1E-05 |  |
|  |  | 70-80% | 936 | 4533 | 1.30 (1.17-1.45) | 6.8E-07 |  |
|  |  | 80-90% | 1031 | 4533 | 1.47 (1.33-1.63) | 2.2E-13 |  |
|  |  | 90-100% | 1069 | 4533 | 1.59 (1.44-1.77) | 4.2E-19 |  |
|  | Chronic Kidney Disease<sup>§</sup> | 0-10% | 3446 | 3391 | 1.00 (Ref) | - | 7.3E-06 |
|  |  | 10-20% | 3490 | 3391 | 1.03 (0.93-1.15) | 0.508 |  |
|  |  | 20-30% | 3439 | 3391 | 1.04 (0.94-1.14) | 0.488 |  |
|  |  | 30-40% | 3463 | 3391 | 1.05 (0.95-1.16) | 0.323 |  |
|  |  | 40-50% | 3370 | 3391 | 1.04 (0.95-1.14) | 0.409 |  |
|  |  | 50-60% | 3362 | 3391 | 1.07 (0.97-1.17) | 0.166 |  |
|  |  | 60-70% | 3389 | 3391 | 1.07 (0.98-1.17) | 0.129 |  |
|  |  | 70-80% | 3285 | 3391 | 1.07 (0.98-1.17) | 0.121 |  |
|  |  | 80-90% | 3373 | 3391 | 1.07 (0.98-1.16) | 0.151 |  |
|  |  | 90-100% | 3326 | 3391 | 1.16 (1.07-1.26) | 5.9E-04 |  |
|  | Neuropathy | 0-10% | 2176 | 3814 | 1.00 (Ref) | - | 7.9E-08 |
|  |  | 10-20% | 2193 | 3814 | 1.03 (0.96-1.11) | 0.436 |  |
|  |  | 20-30% | 2217 | 3814 | 1.07 (0.99-1.15) | 0.075 |  |
|  |  | 30-40% | 2218 | 3814 | 1.06 (0.99-1.15) | 0.110 |  |
|  |  | 40-50% | 2217 | 3814 | 1.05 (0.98-1.13) | 0.192 |  |
|  |  | 50-60% | 2293 | 3814 | 1.11 (1.03-1.20) | 0.006 |  |
|  |  | 60-70% | 2261 | 3814 | 1.10 (1.02-1.18) | 0.014 |  |
|  |  | 70-80% | 2253 | 3814 | 1.10 (1.02-1.19) | 0.009 |  |
|  |  | 80-90% | 2265 | 3814 | 1.11 (1.03-1.19) | 0.007 |  |
|  |  | 90-100% | 2377 | 3814 | 1.21 (1.12-1.30) | 9.7E-07 |  |

**Table 5:** Phewas of 2 LoF variants in MVP whites

| Gene | rsid | AA.change | phewasPhenotype | P | Cases | Controls | OR | lowCI | highCI |
| --- | --- | --- | --- | --- | --- | --- | --- | --- | --- |
| ANKDD1B | rs34358 | p.Trp480* | Diabetes mellitus | 1.04E-06 | 62930 | 104442 | 0.96 | 0.95 | 0.98 |
|  |  |  | Type 2 diabetes | 1.36E-06 | 62531 | 104442 | 0.96 | 0.95 | 0.98 |
|  |  |  | Type 2 diabetes with neurological manifestations | 1.63E-05 | 14159 | 104442 | 0.94 | 0.92 | 0.97 |
|  |  |  | Disorders of lipid metabolism | 5.03E-08 | 141535 | 41406 | 1.05 | 1.03 | 1.07 |
|  |  |  | Hyperlipidemia | 4.66E-08 | 141408 | 41406 | 1.05 | 1.03 | 1.07 |
|  |  |  | Hypercholesterolemia | 2.33E-06 | 32008 | 41406 | 1.06 | 1.03 | 1.08 |
| CCHCR1 | rs3130453 | p.Trp78* | Diabetes mellitus | 4.26E-05 | 62930 | 104442 | 0.97 | 0.96 | 0.98 |
|  |  |  | Type 1 diabetes | 3.99E-07 | 6566 | 104442 | 0.91 | 0.88 | 0.95 |
|  |  |  | Type 2 diabetes | 3.96E-05 | 62531 | 104442 | 0.97 | 0.96 | 0.98 |
|  |  |  | Epistaxis or throat hemorrhage | 1.96E-05 | 2751 | 110902 | 1.12 | 1.07 | 1.19 |
|  |  |  | Celiac disease | 2.72E-19 | 418 | 124470 | 0.52 | 0.45 | 0.60 |
|  |  |  | Microscopic hematuria | 1.83E-05 | 4078 | 147054 | 1.1 | 1.05 | 1.15 |
|  |  |  | Psoriatic arthropathy | 7.82E-10 | 1077 | 140876 | 0.76 | 0.70 | 0.83 |

We evaluated whether the T2D gPRS was associated with the risk of micro- and macrovascular outcomes in an analysis restricted to participants with T2D. The P-value are calculated using gPRS as a continuous exposure and odds ratios are shown by contrasting the top to the bottom gPRS decile (Figure 2 and Table 4a-b). The T2D gPRS was more strongly associated with microvascular than macrovascular outcomes, in particular with retinopathy, but also with neuropathy and CKD. For macrovascular outcomes, T2D gPRS was associated with the risk of PAD, but not with the risk of CHD or acute ischemic stroke.

## Discussion

We report the discovery of 318 novel autosomal and X-chromosome variants associated with T2D susceptibility in a large multi-ethnic T2D study. Furthermore, we observed 13 variants associated with differences in T2D-related micro- and macrovascular outcomes between diabetic and non-diabetics. The substantial locus discovery was achieved by combining data from several large-scale biobanks and consortia, where the MVP data constituted over 40% of all T2D cases. Furthermore, we present the largest cohort of African Americans including over 56K subjects, substantially greater than any African-specific T2D study published to date.

Analyses of coding variants identified 44 variants for T2D, including three pLoF variants in *LPL, ANKDD1B* and *CCHCR1*. 804 putative causal genes at both novel and previously reported loci were identified and 54 genes were found to be possible targets for FDA-approved drugs and chemical compounds. Our SNP-T2D interaction analyses identified several loci where the association between a genetic variant and a vascular outcome differed between people with T2D as compared to those without. We further found that a high polygenic risk for T2D strongly increased the risk for retinopathy in patients with T2D, and also for CKD, neuropathy, and PAD.

T2D is highly prevalent in people of African ancestry; however there are a total of three published T2D GWAS reports in this ancestral group with only 4 definitely detected loci^18,19,20^. In our study with over 56K participants of recent African ancestry, we report 4 novel loci for T2D that are solely observed in this ancestral group, including one that is located on chromosome X. Of the previously reported loci, only rs3842770 (INS-IGF2) was replicated here. The reported HLA-B variant rs2244020^18^ did not replicate in our study; but we did observe a significant association with another SNP in the HLA region (rs10305420, OR 1.15, P = 8.5×10^−9^). We did not replicate the association with the variant rs7560163^20^. Finally, 1 locus (rs73284431) that was recently identified in a large study conducted in Sub-Saharan African countries^19^ failed to replicate in our study consisting of African Americans despite comparable minor allele frequencies (9% vs 10%). Finally, we observed that one *SLC30A8* variant showed a very prominent T2D effect in African Americans that was absent in other populations; mouse studies have shown that the human R138X LoF variant in *SLC30A8* results in increased insulin secretory capacity^21^.

The sole presence of a coding variant near a tagging SNP does not constitute enough evidence to infer a causal association. However, recent exome-array genotyping of over 350K subjects identified 40 coding variants associated with T2D, of which 26 mapped to near known risk-associated loci^22^.

Similarly, an exome sequencing study in over 40K participants reported 15 variants associated with T2D, of which only 2 were not previously reported by GWAS^13^. Sequencing efforts are indispensable for identifying causal variants and genes related to disease, as well as providing insight into the contributions of ultra-rare alleles, but have also substantiated the value of array-based association studies.

Our transcriptome-wide analyses identified 804 putatively causal genes, including 54 genes that appear to be regulated by approved drugs and 687 genes that have not been previously reported. Some of these genes are already well established for T2D etiology (e.g. *KCNJ11*). Except for skeletal muscle, the tissues that showed strongest associations are not known to be of importance in T2D etiology. This raises the hypothesis that the eQTL associations are universal and not tissue dependent; if the gene is expressed in the respective tissue at reasonable levels an association will be detected. We did not observe any significant association in the alpha and beta islet cells, which could be the result of the small sample size (e.g. 30 alpha cells and 19 beta cells). In addition, whole islet transcriptomes are notoriously variable due to the large differences in islet composition among humans, and a few transcripts make up half the transcriptome^23^.

Of particular clinical importance, we identified several genes which are therapeutic targets for medications in patients that are treated for cardiometabolic conditions. We identified two genes, *SCN3A* and *SV2A* whose expression is modified by anti-epileptic agents, and evidence exists showing that anti-epileptic agents may influence glucose regulation. A randomized-controlled trial has reported that the anticonvulsant valproic acid lowers blood glucose concentrations^24^. The information from the gene-drug screen may facilitate future drug repurposing screens.

DEPICT enrichment analysis identified AKT2 subnetwork gene set to be show the highest association with T2D. Of interest is that *AKT2* is critical to insulin signaling and not beta-cell function. The majority of the early era T2D GWAS studies^25^ predominantly identified genetic variants that alter T2D risk through reduced beta-cell function, such as *KCNJ11, HHEX, SLC30A8, CDKAL1, TCF7L2, CDKN2A/2B*, and *IGF2BP2*. However, as the study sample size have substantially increased and the number of T2D-associated variants is approaching the 1.000 number mark, the cumulative effect of genetic variants on T2D is pointing towards a predominant role for insulin resistance.

When we evaluated polygenic risk scores and T2D-related outcomes, we observed that among vascular outcomes, the T2D gPRS was most significantly associated with retinopathy. It is possible that the use of the T2D gPRS provides an opportunity to identify patients who are at the highest risk of developing microvascular complications, such as retinopathy. Using the T2D gPRS, we observed significant associations with other T2D-related outcomes such as CKD, PAD, and neuropathy. Studies at specific loci using both common and rare coding variants will be required to understand pathways leading to T2D-related vascular outcomes.

In a SNP-T2D interactions analysis on T2D-related vascular outcomes we identified 13 loci where the effect on outcome was different between the strata of T2D, of which 3 occurred at previously established variants and 10 had not been previously reported. Our findings have clinical translational potential for risk-stratification and identify diabetic patients who are predisposed to develop subsequent vascular outcomes and present therapeutic opportunities to attenuate the risk of diabetes progression in individuals with T2D.

For T2D-related retinopathy, four variants were found to have different effect sizes between people with and without T2D. The strongest signal for interaction in relation to retinopathy was observed for *GJA8*. Deletion of this gene has been associated with eye abnormalities and retinopathy of prematurity in premature infants, inherited cataracts, visual impairment and cardiac defects and eye abnormalities^26–28^. *TCF7L2* is a known diabetes locus and its association with progression to retinopathy has been previously established^29^. *SLC18A2* is expressed in adult retina and retinal pigment epithelium tissues^30^; this gene is involved in the transport of monoamines into secretory vesicles for exocytosis^31^. *SVILP1* has been previously shown to be associated with thiamine (vitamin B1) prescription, which is frequently prescribed to people with blurry vision^32^.

For chronic kidney disease, two loci, *UMOD* and *TENM3*, were identified for gene-T2D interaction effects. *UMOD* encodes for uromodilin which is exclusively produced by the kidney tubule where it plays an important role in kidney and urine function^33^. A large-scale study in over 133K participants has shown that the serum creatinine-lowering allele in *UMOD* (rs12917707) is more prevalent in diabetic individuals with CKD as compared to diabetic participants without CKD^34^. Variation in *TENM3* has been associated with cholangitis and kidney disorders in the UK Biobank ^35^.

SNP-T2D interaction analysis of neuropathy identified one locus, *NRP2. NRP2* encodes for neuropilin-2 which is an essential cell surface receptor involved in VEGF-dependent angiogenesis and sensory nerve regeneration^36,37^.

For coronary heart disease, we identified several SNP-T2D interactions. Variation at 9p21 has been reported previously for CHD and T2D. *SORT1* is a lipid-associated locus; in our analyses, allelic variation at this locus that decreases CHD risk and decreases lipids conferred a stronger protection in people with T2D compared to those without T2D. Coupled with findings in mice that identified *SORT1* as a novel target of insulin signaling^38^, our findings raise the hypothesis that *SORT1* may contribute to altered hepatic apoB metabolism under insulin-resistant conditions.

The SNP rs71039916 is located nearby *PDE3A*, and collocates with a SNP (rs3752728, D’ = 0.867, r^2^ = 0.08) that is associated with diastolic blood pressure^39,40^. As a phosphodiesterase that reduces cAMP levels, the PDE3A protein limits protein kinase A/cAMP signaling, and has been shown to affect proliferation of vascular smooth muscle cells^41^. Cell line research has shown that cAMP levels might impact the regulation of insulin secretion in pancreatic β-cells^42^, and more recent gene ablation studies in mice have established that cAMP/CREB signaling controls the insulinotropic and anti-apoptotic effects of GLP-1 signaling in adult mouse β-cells^43^. Subcutaneous adipose tissue of patients with T2D show increased PDE activity, and inverse correlations between total PDE3 activity and BMI have been reported in adipocytes^44^.

We have conducted the largest discovery study for T2D to date, including over 220K T2D cases and 1.2 million controls. Over 21% of our discovery sample comprised of non-European participants; our African American sample included over 56K subjects. A unique strength in our study is the comprehensive individual level phenotype data available for analyzing T2D and related micro- and macrovascular outcomes. This allowed us to test for genetic interaction instead of evaluating effect differences between strata using summary statistics.

In summary, we have identified 318 novel genetic variants associated with T2D risk and T2D-related vascular outcomes, including 3 population-specific autosomal loci in African Americans, 8 chromosome-X variants, plus an additional 13 variants associated with differences in T2D-related micro- and macrovascular outcomes across diabetic stratum. Coding variant analyses and transcriptome wide analyses have identified many putative causal genes that could be pursued in functional mechanistic studies and provide an updated map of effects for gene-tissue pairs for T2D risk. Our gene-drug analyses provide a genetic basis to guide future drug repurposing studies. Lastly, our SNP-T2D interaction analyses elucidate differences in pathogenetic mechanisms of T2D-related vascular outcomes with potential clinical implications for preventing development of microvascular complications in T2D.

## Online Methods

We conducted a large-scale multi-ethnic autosomal and X-chromosome T2D GWAS of common variants in over 1.4 million participants. We subsequently conducted analyses to facilitate the prioritization of these individual findings, including transcriptome-wide predicted gene expression for T2D, secondary signal analysis, T2D-related vascular outcomes analysis using SNP-T2D interaction analysis, coding variant mapping, phenome-wide association analyses of pLoF variants, and a drug repurposing screen.

### Discovery cohort

The Million Veteran Program (MVP) is a large cohort of fully consented veterans of the United States military forces recruited from 63 participating Department of Veterans Affairs (VA) medical facilities^3^. Recruitment started in 2011, and all veterans were eligible for participation. Compared with the general population, the Million Veteran Program is overrepresented with participants of male gender, but the study is representative of the US population in terms of race and ethnicity. This study analyzed clinical data through July 2017 for participants who enrolled between January 2011 and October 2016. Across ancestries, the average age at study enrollment ranged from 56.1 for Asian to 68.2 for European participants (Supplementary Table 3). Average body mass index (BMI) ranged from 28.5 for Asians to 30.8 for African Americans. The proportion of males ranged from 87.2% for African Americans to 93.9% for Asians. The prevalence of T2D was 35.5% for Europeans, 36.4% for Asians, 42.1% for Hispanics, and 43.6% for African Americans. All MVP study participants provided blood samples for DNA extraction and genotyping, and completed surveys about their health, lifestyle, and military experiences. Consent to participate and permission to re-contact was provided after counseling by research staff and mailing of informational materials. Study participation includes consenting to access to the participant’s electronic health records for research purposes, data that captures a median follow-up time of 10.0 years at time of study enrollment. Each veteran’s electronic health care record is integrated into the MVP biorepository, including inpatient International Classification of Diseases (ICD-9-CM and ICD-10-CM) diagnosis codes, Current Procedural Terminology (CPT) procedure codes, clinical laboratory measurements, and reports of diagnostic imaging modalities. Researchers are provided data that is de-identified except for dates, and do not have the ability or authorization to link these details with a participant’s identity. Blood samples are collected by phlebotomists and banked at the VA Central Biorepository in Boston, where DNA is extracted and shipped to two external centers for genotyping. The Million Veteran Program received ethical and study protocol approval from the VA Central Institutional Review Board (cIRB) in accordance with the principles outlined in the Declaration of Helsinki.

#### Genotyping

DNA extracted from buffy coat was genotyped using a custom Affymetrix Axiom biobank array. The MVP 1.0 genotyping array contains a total of 723,305 SNPs, enriched for 1) low frequency variants in African and Hispanic populations, and 2) variants associated with diseases common to the VA population^3^.

#### Genotype quality-control

The MVP genomics working group applied standard quality control and genotype calling algorithms to the data in two batches using the Affymetrix Power Tools Suite (v1.18). Standard quality control pipelines were used to exclude duplicate samples, samples with more heterozygosity than expected, samples with an excess (> 2.5%) of missing genotype calls and samples with discordance of genetically inferred sex versus self-report. We excluded related individuals (halfway between second- and third-degree relatives or closer) as measured by the KING software^45^. Before imputation, variants that were poorly called or that deviated from their expected allele frequency based on reference data from the 1000 Genomes Project were excluded ^46^. After prephasing using EAGLE v2^47^, genotypes from the 1000 Genomes Project phase 3, version 5 reference panel were imputed into MVP participants via Minimac3 software^48^. The top 30 principal components were computed using FlashPCA^49^ on a genotype dataset that included all MVP participants and an additional 2,504 individuals from 1000 Genomes. These 30 PCs were then used for the unification of self-reported race/ancestry and genetically inferred ancestry to compose ancestral groups used for stratification in subsequent association analyses^50^.

#### Race and ethnicity

Information on race (Europeans, African Americans, Asians and Native Americans) and ethnicity (Hispanic, yes or no) was obtained based on self-report through centralized VA data collection methods using standardized survey forms, or through the use of information from the VA Corporate Data Warehouse or Observational Medical Outcomes Partnership data, when information from self-report survey was missing. Even though self-reported race/ethnicity was missing in 3.67% of study participants, 39.4% of participants had some form of discordant information between the various sources of which the data was extracted. Race and ethnicity categories were merged to form the following race or ancestral groups: Europeans, African Americans, Hispanics, and Asians using a unifying classification algorithm based on self-identified race/ethnicity and genetically inferred ancestral information, termed HARE (Harmonized Ancestry and Race/Ethnicity)^50^. Using this approach, of the total of 351,820 genotyped individuals all but 6,257 (1.78%) were assigned to one of the four ancestral groups.

#### Phenotype classification

ICD-9-CM diagnosis codes from electronic health care records were available for MVP participants from as early as 1998. Participants were classified as a T2D case if they had 2 or more T2D-related diagnosis codes (ICD-9-CM 250.2x) from VA or fee basis inpatient stays or face-to-face primary care outpatient visits in the 731 days before the enrollment date up to July 1^st^ of 2017, excluding those with co-occurring diagnosis codes for T1D (250.1x), secondary or other diabetes or a medical condition that may cause diabetes (249.xx). Participants were selected as controls if they had no ICD-9-CM diagnosis code for type 1, type 2, or secondary diabetes mellitus up to July 2017.

For T2D-related vascular outcomes, the following definitions were used: CHD, at least one admission to a VA hospital with discharge diagnosis of admission for myocardial information, or at least one procedure code for revascularization (coronary artery bypass grafting, percutaneous coronary intervention), or at least 2 ICD-9-CM codes for CAD (410 to 414) registered on at least 2 separate encounters. Subjects were classified as having PAD if they had at least two of ICD-9-CM codes or CPT codes as outlined in the report of Klarin *et al*.^14^, or having one code and at least two visits to a vascular surgeon within a 14 month period. Acute ischemic stroke was defined if at least 1 ICD-9-CM discharge diagnosis code for stroke excluding head injury or rehab (433.x1, 434 [excluding 434.x0], and 436) was present^51^. CKD was classified as an estimated glomerular filtration rate <60 mL/min−1·1.73 m−2 on 2 separate measurements 90 days apart, or ICD-9-CM diagnosis codes for chronic renal failure (585) and/or a history of kidney transplantation (ICD-9-CM V42)^52^; Neuropathy was defined using the following ICD-9-CM diagnosis codes: diabetic neuropathy (356.9, 250.6), amyotrophy (358.1), cranial nerve palsy (951.0, 951.1, 951.3), mono-neuropathy (354.0-355.9), Charcot’s arthropathy (713.5), polyneuropathy (357.2), neurogenic bladder (596.54), autonomic neuropathy (337.0, 337.1), orthostatic hypotension (458). Retinopathy was defined using ICD-9-DM diagnosis codes for: T2D with ophthalmic manifestations (250.50, 250.52), retinal detachments and defects (361.0, 361.1), disorders of vitreous body (379.2), other retinal disorders (362.0, 362.1, 362.3, 362.81, 362.83, 362.84), excluding ICD-9-CM codes associated with macular degeneration (362.5).

#### MVP analysis

We tested imputed SNPs that passed quality control (e.g. HWE > 1.0×10^−10^, INFO > 0.3, call rate > 0.975) for association with T2D through logistic regression assuming an additive model of variants with MAF > 0.1% in Europeans, and MAF > 1% in African Americans, Hispanics and Asians using PLINK2a software^53^. Covariates included age, gender, and 10 principal components of genetic ancestry.

Other studies: Summary statistics available from previous reports in the DIAMANTE Consortium^4^, Biobank Japan^7^, Penn Medicine Biobank^5^, Malmö Diet and Cancer Study^8^, MedStar/PennCath studies^9^, and the Pakistani Genomic Resource^6^ were obtained for meta-analysis (Supplemental Table 2). All cohort were imputed using the 1000 Genomes Project phase 3, version 5 reference panel with exception of the DIAMANTE consortium where genotype calls were imputed using the Haplotype Reference Consortium reference panel^54^. Briefly, following central and study-specific quality control protocols, an additional 917,392 Europeans, 2,647 African Americans, and 213,834 Asians were available for T2D meta-analysis. Only SNPs with ancestry-specific MAF > 1% in these studies were used. Within each study a logistic regression model was used where T2D was set as the dependent variable, imputed SNP as the independent variable, and analyses were adjusted for age, gender, and the top ten principal components of genetic ancestry.

#### Meta-analysis

We aggregated association summary statistics from the above mentioned studies to perform ancestry-specific and multi-ethnic meta-analysis. The association summary statistics for each analysis were meta-analyzed in a fixed-effects model using METAL with inverse-variance weighting of log odds ratios^55^. From the meta-analysis, all variants were extracted that passed quality control.

Between-study allelic effect size heterogeneity was assessed with Cochran’s Q statistic as implemented in METAL. Variants were considered genome-wide significant if they passed the conventional p-value threshold of 5.0×10-8. We excluded variants with a high amount of heterogeneity (I^2^ statistic > 75%) across the four evaluated ancestral groups.

#### Novel loci

Novel loci were defined as those in which the lead variant was not in LD or physically nearby (500 Kb) to a previously reported lead GWAS variant(s). For each locus, we first searched ± 500 Kb surrounding the lead SNP to ensure that potential long-range genetic influences were assessed.

#### Chromosome-X analysis

X chromosome genotypes were processed separately from autosomal genotypes. During prephasing and imputation an additional flag of -chrX was added. In post-imputation stage, the XWAS QC pipeline was employed to remove variants in the pseudo-autosomal regions, variants that were not in HWE in females (P > 1.0×10^−6^), variants with differential allele frequencies, and variants with differential missingness (p<10−7) between males and female controls (Supplemental Figure 2)^56^. For each ancestry-specific subset, we performed sex-stratified analysis where dosages (number of chromosome copies) for the X-chromosome in T2D cases are equivalent to controls within each sex stratum. For the meta-analysis of sexes, differences in dosage compensation should be considered between males (with one X chromosome) and females (with two X chromosomes). In a separate verification analysis we performed a chromosome-X association analysis on dosage data using PLINK (--xchr-model) which produced consistent results compared with analysis of best-guess data where the X chromosome was coded as 0/2 for males instead of 0/1. The output from sex-stratified chromosome-X analyses were first meta-analyzed into a multi-ethnic sex-stratified analysis. Then, the multi-ethnic results from males and multi-ethnic results from females were meta-analyzed, where none of the analyzed variants was detected using the Cochrane test for heterogeneity (P < 5.0×10^−8^). Results presented include overall effect estimates, male-specific effect estimates, female-specific estimates, and the heterogeneity p-value (Supplementary Table 13).

### Secondary signal analysis

GCTA^57^ was used to conduct conditional analyses to detect ancestry-specific distinct association signals at each of the lead SNPs utilizing the meta-analyzed GWAS summary statistics in Europeans and African Americans. Race-stratified MVP cohorts (197,066 Europeans and 53,445 African Americans) were used to model LD patterns between variants. The reference panel of genotypes consisted of the variants with allele frequencies > 0.1% in Europeans and >1% in African Americans that passed quality control criteria in the MVP-specific T2D GWAS (INFO > 0.3, HWE P > 1.0×10^−10^, call rate > 0.975). For each lead SNP, conditionally independent variants that reached locus-wide significance (P < 1.0×10^−5^) were considered as secondary signals of distinct association. If the minimum distance between any distinct signals from two separate loci was less than 500 Kb, we performed additional conditional analysis including both regions and reassessed the independence of each signal. Finally, the predicted conditionally independent variants were tested in a logistic regression model in the MVP study only to empirically validate the signal, and results are shown in Supplemental Tables 7 and 10.

### Coding variant mapping

All imputed variants in MVP were evaluated with Ensemble variant effect predictor^58^, and all predicted LoF and missense variants were extracted. The LD was calculated with established variants, and in a second round of analysis the effect of the missense variant was calculated conditioning on the lead SNP to assess how much residual variance the SNP explains in T2D risk. A p-value of 0.05 was considered as statistically significant.

### S-PrediXcan and colocalization analyses

Genetically predicted gene expression and its association with T2D risk was estimated using S-PrediXcan^15^. Input included meta-analyzed summary statistics from the European T2D GWAS for common variants and reference eQTL summary statistics for 52 tissues including 48 tissues from GTEx^59^, 2 cell types in kidney tissue (glomerulus and tubulus)^60^, and 2 cell types in pancreatic islet tissue (alpha and beta)^61^. Analyses incorporated genotype covariance matrices based on 1000 Genomes^46^ European populations to account for LD structure. Colocalization analysis was performed to address the issue of LD-contamination in S-PrediXcan analyses^62^. Input data were identical to those evaluated by S-PrediXcan and colocalization was restricted to only variants included in the S-PrediXcan gene^63^. The output is shown in Supplemental Table 8.

### Phenotypic variance explained by SNPs

The MVP European cohort (69,869 T2D cases and 127,197 controls) was used to calculate the variance explained by the 558 genome-wide-significant sentinel SNPs from the multi-ethnic meta-analysis. A logistic regression model was performed assuming additive effects where T2D was set as the dependent variable, and the sentinel SNPs as independent variables, adjusting for sex, age, and 10 principal components. The baseline model included age, gender and 10 principal components. The variance explained was calculated with the r^2^ statistic, calculated as 1 – (residual deviance / null deviance).

### Phenome-wide association analysis

For the three LoF variants that were identified using coding variant analysis, we performed a PheWAS study^64^ to fully leverage the diverse nature of the Million Veteran Program as well as the full catalog of relevant ICD-9-CM diagnosis and CPT procedure codes (Supplementary Table 18). Of genotyped veterans, participants were included in the PheWAS analysis if their respective electronic health record reflected two or more separate encounters in the VA Healthcare System in each of the two years prior to enrollment in MVP. Using this method, a total of 277,531 veterans spanning 21,209,658 available ICD-9 diagnosis codes were available for PheWAS analysis. We restricted our analysis on the subgroup of 197,066 European participants. Diagnosis and procedure codes were collapsed to clinical disease groups and corresponding controls using predefined groupings^65^. Phenotypes were required to have a case count over 25 in order to be included in the PheWAS analysis, and a multiple testing thresholds for statistical significance was set to P < 2.8×10^−5^ (Bonferroni method). Each of the previously unpublished LoF variants (ANKDD1B p.Trp480* and CCHCR1 p.Trp78*) were tested using logistic regression adjusting for age, sex, and 10 principal components in an additive effects model using the PheWAS R package in R v3.2.0^66^. The results from these analyses are reported as odds ratios, in which the estimate is the average change in odds of the PheWAS trait per weighted T2D-increasing allele (Supplementary Table 18, Supplementary Figure 4).

### Analysis of T2D-related outcomes

Genetic data on European participants was separately analyzed using outcomes as a binary outcome, and T2D as an interaction variable with SNPs using interaction analysis with robust variance to reduce effect heteroscedacity (e.g. unequal variance between strata) using SUGEN software (v8.8)^67^. We evaluated the interaction between SNP and presence of T2D status using an interaction term for the two independent variables. Due to the binary nature of the outcome, the standard output from the interaction effect estimate are interpreted on a multiplicative scale, e.g., the departure from the product of the two independent effects. To obtain interaction on an additive scale (e.g. departure from the sum of the independent effects), we calculated the relative excess risk due to interaction (RERI) metric, a well-known analytic method in classic epidemiologic literature^68^. In case-control studies using the linear additive odds-ratio model as proposed by Richardson and Kaufman^68^ in our study has the form of:

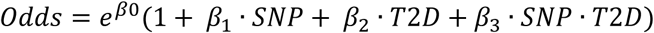

In which the coefficient *β*_3_ measures the departure from additivity of exposure effect on an odds ratio scale; that is

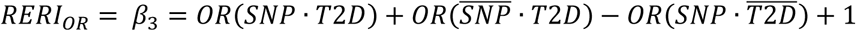

Where 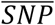 and 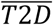 note the absence of the respective covariates.

In contrast to the standard exponential odds model, where the effects of explanatory variables are additive on an exponential scale, we performed analysis using a linear odds model, in order to quantify the excess odds per unit of the given explanatory variables on the outcome. In this model, the excess relative risk due to interaction is an estimate of the excess odds on a linear scale due to the interaction between two explanatory variables (in our case, T2D state and genotype dosage of the tested variant). In the SNPxT2D interaction analysis we used a significance threshold of P < 5.0×10^−08^ to denote variants that statistically different effect sizes between participants with T2D as compared those without. An additional filter was applied, and variants for which the effect size in at least one of the two T2D strata was nominally significant at P < 0.001 were included. Manhattan plots and the table are used to represent the interaction coefficients on this scale.

### Polygenic risk scores and risk of T2D and related outcomes

We constructed a genome-wide polygenic risk score (gPRS) for T2D in the MVP participants of European ancestry by calculating a linear combination of weights derived from the Europeans in the DIAMANTE Consortium^4^ using the prune and threshold method in PRSice software^69^. After an initial sensitivity analysis, the r^2^ threshold for pruning was set to 0.8, and the P-value for significance threshold was set to 0.05. The gPRSs were divided into deciles and the risk of T2D was assessed using a logistic regression model using the lowest decile as a reference (e.g. the 10% of participants with lowest of T2D gPRS), together with the potential confounding factors of age, gender, BMI, and the first 10 principal components of European ancestry. An additional outcomes analysis was performed to evaluate to what extent a T2D gPRS is predictive of T2D-induced morbidities. The dataset was restricted to subjects with T2D, and a stratum-restricted T2D gPRS deciles were generated. Logistic regression models were applied where the micro- and macrovascular conditions were modeled as outcomes, and independent variables included strata-restricted gPRS deciles, age, gender, and the first 10 principal components of European ancestry. The data were visualized using shape-plots.

### Heritability estimates and genetic correlations with other complex traits and diseases

LD-score regression was used to estimate the heritability coefficient, and subsequently population and sample prevalence estimates were applied to estimate heritability on the liability scale^70^. A genome-wide genetic correlation analysis was performed to investigate possible co-regulation or a shared genetic basis between T2D and other complex traits and diseases. Pairwise genetic correlation coefficients were estimated between the meta-analyzed T2D GWAS summary output in Europeans and each of 774 precomputed and publicly available GWAS summary statistics for complex traits and diseases by using LD score regression through LD Hub v1.9.3 (http://ldsc.broadinstitute.org). Statistical significance was set to a Bonferroni-corrected level of P < 6.5×10^−5^.

### Enrichment and pathway analyses

Tissue enrichment for S-PrediXcan results was evaluated by calculating exact P-values for under- or over-enrichment based on the cumulative distribution function of the hypergeometric distribution. The Bonferroni-corrected threshold for significance was P < 0.001 considering evaluation of 52 tissues.

Enrichment analyses in DEPICT ^71^ were conducted using genome-wide significant (P < 5.0×10^−8^) T2D GWAS lead SNPs. DEPICT is based on predefined phenotypic gene sets from multiple databases and Affymetrix HGU133a2.0 expression microarray data from >37k subjects to build highly-expressed gene sets for Medical Subject Heading (MeSH) tissue and cell type annotations. Output includes a P-value for enrichment and a yes/no indicator of whether the FDR q-value is significant (P < 0.05). Tissue and gene-set enrichment features were considered.

### Evaluation of drug classes for genes with associations with gene expression

To identify drug-gene pairs that may be leads for repurposing or may be attractive leads for novel inhibitory drugs, we identified drugs targeting genes whose predicted expression was significantly associated with T2D risk in S-PrediXcan analyses and which we predicted would lower blood glucose based on direction of effect on T2D risk with increasing gene expression and drug action (activator or inhibitor). Medications with a primary indication for diabetes and medications with adverse drug events for diabetic patients were evaluated using the SIDe Effect Resource (SIDER)^16^. Medications targeting genes were queried using DGIdb^17^. These drug targets represent a set of genes that are both likely to be involved in glucose regulation in one or more tissues and can be targeted by drugs. Genes and medications identified in this analysis are presented in Supplementary Table 14.

## Data Availability

The full summary level association data from the trans-ancestry meta-analysis from this report will be available through dbGAP (accession codes will be available before publication).

## Acknowledgements

This research is based on data from the Million Veteran Program, Office of Research and Development, Veterans Health Administration and was supported by award no. MVP000. This publication does not represent the views of the Department of Veterans Affairs, the US Food and Drug Administration, or the US Government. This research was also supported by funding from: the Department of Veterans Affairs award I01-BX003362 (P.S.T. and K.M.C) and the VA Informatics and Computing Infrastructure (VINCI) VA HSR RES 130457 (S.L.D) B.F.V. acknowledges support for this work from the NIH/NIDDK (DK101478), the NIH/NHGRI (HG010067) and a Linda Pechenik Montague Investigator award. K.M.C, S.M.D, J.M.G, C.J.O, L.S.P, and P.S.T. are supported by the VA Cooperative Studies Program. S.M.D. is supported by the Veterans Administration [IK2-CX001780]. D.K. is supported by the National Heart, Lung, and Blood Institute of the National Institutes of Health [T32 HL007734]. L.S.P. is supported in part by VA awards I01-CX001025, and I01CX001737, NIH awards R21DK099716, U01 DK091958, U01 DK098246, P30DK111024, and R03AI133172, and a Cystic Fibrosis Foundation award PHILLI12A0. We thank all study participants for their contribution. Data on T2D have been contributed by investigators from DIAMANTE Consortium, Biobank Japan, Malmö Diet and Cancer Study, PennCath, MedStar, Pakistan Genomic Resource, Penn Medicine Biobank, and Regeneron Genetics Center. Data on stroke were provided by MEGASTROKE investigators, and data on CKD have been contributed by CKDgen investigators. Data on alpha and beta islet cells have been contributed by the HPAP Consortium. Data on coronary artery disease have been contributed by the CARDIoGRAMplusC4D investigators.

## Ethics statement

The Central Veterans Affairs Institutional Review Board (IRB) and site-specific Research and Development Committees approved the Million Veteran Program study. The Vanderbilt University Medical Center IRB approved the use of BioVU data for this study. All other cohorts participating in this meta-analysis have ethical approval from their local institutions. All relevant ethical regulations were followed.

## Consortia

VA Million Veteran Program

Regeneron Genetics Center

The HPAP Consortium

## Contributions

M.V., J.M.K., K.-M.C., D.S., B.F.V., P.S.T., C.J.O. were responsible for the concept and design. The acquisition, analysis or interpretation of data were performed by M.V., J.M.K., K.-M.C., D.S., B.F.V., P.S.T., R.L.J., C.T., T.L.A., J.E.H., J.Z., J.H., K.M.L, D.K., S.P., J.D., O.M., A.R., N.Q., S.S.S., S.H., I.H.Q, M.N.A, U.J., A.J., S.A., X.S., L.G., K.K., K.S., Y.V.S., S.L.D., K.C., J.S.L., J.M.G., L.S.P., D.R.M., J.B.M., P.R., P.W.W., T.L.E., D.J.R., S.M.D., and C.J.O. The authors M.V. and D.S. drafted the manuscript. The critical revision of the manuscript for important intellectual content was carried out by M.V., J.M.K., K.-M.C., D.S., B.F.V., P.S.T., C.T., J.Z., J.H., D.K., X.S., L.G., K.K., K.S., L.S.P., J.B.M., P.R., T.L.E., S.M.D., and C.J.O. Finally, K.-M.C., D.S., and B.F.V. provided administrative, technical, or material support.

## Reporting Summary

Further information on research design is available in the Nature Research Reporting Summary linked to this article.

## Data availability

The full summary level association data from the trans-ancestry, European, African American, Hispanic, and Asian meta-analysis from this report will be available through dbGAP (Accession codes will be available before publication).

## Code availability

Imputation was performed using MiniMac4 and EAGLE, association analysis was performed using PLINK2A and XWAS. Post-GWAS processing software include: FlashPCA, KING, METAL, GCTA-COJO, S-PrediXcan, SUGEN, DEPICT, SIDER, and DGidb as outlined in the Online Methods. Clear code for analysis is available at their associated websites. Additional analyses were performed in R-3.2.

